# Long-read genome sequencing resolves complex structural variants in exome-negative developmental and epileptic encephalopathy

**DOI:** 10.64898/2026.10.26.26365902

**Authors:** Joohyun Park, Zakaria Eddafir, Josua Kegele, Davide Mei, Noor Smal, Christina Vossler-Wolf, Kornelia Ellwanger, Charissa Millevert, Merel Swinnen, Robert Lauerer-Braun, Heidi Löffler, Thomas Ott, German Demidov, Michael Alber, Francesca Bisulli, Susan Byrne, Laura Canafoglia, Evelina Carapancea, Barbara Castellotti, Gianpiero Cavalleri, Nicolas Chatron, John Coleman, Maria Roberta Cilio, Tim de Pooter, Peter De Rijk, Wouter De Coster, Els De Vriendt, Matthias De Wachter, Anna Esteve-Codina, Flavia Fineschi, Elena Freri, Julien Gagneur, Hamidah Ghani, Mandana Gheisari, Anna Jansen, Janna Kenny, Roland Krause, Peter Krieg, Laura Licchetta, Oliver Maier, Eva Matzker, Raffaella Minardi, Hiltrud Muhle, Bernd A. Neubauer, Simone Olivieri, Stephan Ossowski, Elena Parrini, Manuela Pendziwiat, Johanna Pohl, Andrea Rüegger, Isabelle Sabatier, Iciar Sanchez-Albisua, Damien Sanlaville, An-Sofie Schoonjans, Anne Schulz, Leon Schütz, Miriam Scida, Hannah Stamberger, Mojca Strazisar, Leo Strothmann, Marc Sturm, Amelie T. van der Ven, Dorothée Ville, Christiane Wagner, Markus Wolff, Carsten Wurst, Vicente A. Yépez, Solve-RD DITF Epicare, Christel Depienne, Renzo Guerrini, Patrick May, Tejasvi Niranjan, Simona Balestrini, Gaëtan Lesca, Tobias B. Haack, Sarah Weckhuysen, Holger Lerche

## Abstract

**Background and Objectives:** Despite standard genetic testing, approximately two-thirds of patients with developmental and epileptic encephalopathy (DEE) remain undiagnosed. Pathogenic noncoding and structural variants (SVs), that are difficult to detect or resolve using short-read sequencing, may contribute to this diagnostic gap. To evaluate the diagnostic contribution of long-read genome sequencing (lrGS), we recruited a cohort of 124 families with sporadic DEE who remained unsolved by prior ES.

**Methods:** Trio lrGS (Oxford Nanopore or PacBio HiFi) was performed, complemented by short-read genome sequencing (srGS) and mRNA sequencing (blood or fibroblast) of index cases, where available. Comprehensive variant analysis included small nucleotide variants, indels, SVs, and short tandem repeats. To identify the diagnostic yield of each method, we compared lrGS, srGS and ES re-analysis, and evaluated the additional value of RNA sequencing.

**Results:** We established a diagnosis in 21 of 124 families (16.9%). Fourteen variants were retrospectively detectable in prior ES. Two cases with non-coding variants in *SCN1A* and *RNU2-2* were detectable by both srGS and lrGS. Five SVs also required genome sequencing (5.6% of the total cohort), three of which could be detected, but not confidently resolved by srGS due to breakpoint complexity, repeat-mediated rearrangements, and large copy-neutral events: LrGS enabled accurate characterization of Alu-mediated *CDKL5* and *STXBP1* deletions and a *MEF2C* deletion with large inversion. A copy-neutral *SCN1A* inversion was well characterizable by both srGS and lrGS. One balanced translocation t(X;17) disrupting *CDKL5* was resolved by lrGS (srGS unavailable). RNA sequencing confirmed pathogenicity of two SVs including t(X;17), the *MEF2C* deletion with inversion, and two CNVs. In addition, *ZNF569* and *ZNF570* emerged as candidate epilepsy genes from our cohort.

**Discussion:** In ES-negative DEE, the incremental diagnostic yield of genome sequencing was modest, while lrGS provided particular value for the structural resolution of complex rearrangements, especially those involving repetitive sequences. The finding that two-thirds of diagnoses were retrospectively detectable in prior ES highlights the substantial value of systematic reanalysis. Further gains from genome sequencing may increasingly depend on improved interpretation of non-coding variation and integration of long-read-specific information. Our findings indicate that complex SVs represent the class for which lrGS currently yields the most distinctive diagnostic information.

## 1. Introduction

Developmental and epileptic encephalopathies (DEEs) are a group of severe neurodevelopmental disorders characterized by often drug-resistant epilepsy, accompanied by varying degrees of developmental impairments.^1^ They span clinically recognisable syndromes such as Dravet syndrome, Lennox-Gastaut syndrome, and infantile epileptic spasms syndrome.^2^ While each syndrome is individually rare, their combined incidence is approximately 1 in 590 children.^3^ Antiseizure medications reduce seizure frequency in only a subset of patients, leaving most children with severe seizures and profound developmental impairment, placing lasting burdens on families and caregivers.^4^ Most DEEs have a genetic etiology.^5^ Identifying this underlying cause can inform parents about prognosis, expected comorbidities, and recurrence risk in future pregnancies, and can increasingly guide selection of targeted therapies.^1^ Genetic testing currently relies on a combination of exome sequencing (ES) and chromosomal microarray testing (CMA), yet these establish a diagnosis in fewer than half of patients, leaving the majority of children with DEE unsolved.^6^ Several factors contribute to this diagnostic gap. First, pathogenic variants may reside in non-coding regions that are not captured by ES.^7^ Secondly, complex structural variants (SVs) and balanced rearrangements contributing to disease aetiology go undetected by CMA.^8^ Short-read genome sequencing (srGS) addresses some of these gaps, increasing yield by approximately 7%,^9^ nevertheless remains limited for detection of large insertions, inversions, repeat-mediated and complex SVs.^10^

Long-read genome sequencing (lrGS) addresses several of these limitations by generating reads that span entire structural variant loci and providing improved read alignment in complex regions, substantially enhancing SV discovery.^11,12^ One recent study increased the diagnostic yield by approximately 10% in a cohort of patients with rare disease using lrGS.^13^ Since DEEs are genetically heterogeneous and may involve complex variants that are typically missed with srGS, they are well suited for lrGS approaches.^14^ However, the clinical utility of lrGS in genetically unresolved DEE remains incompletely defined, particularly regarding which diagnostic findings genuinely require long-read sequencing versus systematic reanalysis of existing ES or short-read genome data.

Here, we applied trio lrGS to 124 families with DEE unsolved after ES, complemented by index srGS and, where available, index RNA sequencing (RNAseq). We evaluated the diagnostic contribution of lrGS across variant classes and specifically assessed whether identified variants could be detected or resolved using available ES and srGS data. Using two in-house developed variant detection pipelines, we demonstrate that trio-based lrGS can uncover clinically relevant variants across multiple variant classes, while providing particular added value for the detection and resolution of complex SVs that may remain difficult to characterize with conventional short-read approaches.

## 2. Methods

### Cohort description

For this collaborative European study, patients with DEE who remained without a molecular diagnosis after ES were selected for lrGS by clinical experts at participating centers. A prior ES was performed either through local diagnostic services or the Epi25 collaborative.^15^ Onset of seizures occurred in the first year of life in 86 individuals, between 1 and 2 years in 32 individuals, and between 2 and 7 years in 6 individuals. The final cohort comprised 121 trios, 2 quartets, and 1 duo of DEE index patients and unaffected parents, recruited from February 2022 onwards.

### Sequencing and data processing

Fresh EDTA blood samples were obtained from all index patients and parents, and genomic DNA was extracted for sequencing. Trio ONT lrGS was performed for 101 families (98 trios, 2 quartets, 1 duo) on the Oxford Nanopore PromethION 24 platform, achieving an average coverage of 32±18x (R10 flow cells 37±19x, R9 21±9x). The remaining 23 trios were contributed through the Solve-RD consortium and sequenced with PacBio HiFi (∼10x coverage; eMethods 1).^16^ Library kits and flow-cell chemistries for both platforms are provided in eMethods 2. SrGS was additionally available for 109 index cases (86 in the ONT cohort and all 23 PacBio trios;). Additionally, RNAseq was available for 97 index cases, comprising 80 samples from whole blood (PAXgene tubes) and 17 from cultured skin-derived fibroblasts, details provided in eMethods 3.

Reads were aligned to GRCh38/hg38 using minimap2^17^ and processed with the standardized internal workflows megSAP or PhAnTOM-LR. small sequence variants were called with Clair3 or Clair3-Nova (depending on the processing site),^18^ SVs with Sniffles2 (v2.4 or v2.6; pipeline-specific implementations in),^19^ CNVs from long-read depth with ClinCNV (10 kb bins),^20^ and repeat expansions with Straglr, focusing on known pathogenic tandem-repeat loci curated by STRchive;^21,22^ reads were phased with longphase or WhatsHap.^23,24^ Because SV-caller version affected sensitivity, every candidate SV was manually curated in IGV verifying breakpoint orientation, additional breakpoints, and mobile-element involvement, and excluding alignment artefacts before classification. An average read-length N50 of 14,963±3,256 bp was reached, with approximately 26,715±3,311 SV calls per sample. Processed datasets were shared and uploaded to the Solve-RD platform RD-connect for collaborative analyses.^25^

### Variant annotation and prioritization

Detected DNA variants were filtered and prioritized based on variant type, variant quality, minor allele frequency (MAF) in population databases, pathogenicity prediction scores, and phenotypic compatibility. MAF thresholds of <0.1% were applied for presumed autosomal dominant (AD) and <1% for autosomal recessive (AR) inheritance. Variants frequently observed in internal cohorts or 1kG-ONT datasets were excluded, whereby retained candidate variants were restricted to ≤3 for AD and ≤10 for AR inheritance models. X-linked variants were filtered according to proband sex and corresponding inheritance patterns. For genes associated with AD disorders, *de novo* variants were prioritized, and for genes associated with AR disorders, homozygous and compound-heterozygous variants were considered. Variants passing population and inheritance-based filtering were further prioritized based on their presence in or near (2kb up-/down-stream) genes from a curated list of established epilepsy and intellectual disability (Epi/ID) genes, and genes associated to the proband’s HPO terms. SVs affecting coding regions and transcription start sites were prioritized as potentially high-impact. SNVs were prioritized if they had CADD scores >20, ClinVar pathogenic/likely pathogenic/uncertain significance annotations, or AlphaMissense scores >0.5. Intronic variants were evaluated using SpliceAI, and variants with a score >0.5 were prioritized.^26,27^ Prioritized variants matching the clinical phenotype were classified according to the American College of Medical Genetics and Genomics (ACMG) and Association for Molecular Pathology (AMP) developed guidelines.^28,29^ Trio-lrGS datasets were analyzed first to evaluate whether diagnostically relevant variants across different variant classes could be reliably identified using lrGS data. Where available, singleton srGS datasets were analyzed in parallel or subsequently by independent researchers, to assess whether (i) variants identified by lrGS were also detectable and adequately characterizable using srGS, and (ii) independent srGS analysis would identify additional potentially pathogenic variants missed by lrGS. Prior ES data were retrospectively reviewed for all diagnostic variants where available. RNAseq was used to assess transcript-level consequences and support variant interpretation. SrGS and RNAseq were not available for all participants.

### Minigene Assay

The splicing impact of the deep-intronic *SCN1A* variant (c.265-2028C>G) was assessed with a pET01 minigene splice assay in HEK293 cells; the complete protocol is provided in eMethods 4.

## 3. Results

We performed lrGS in 124 trios with DEE previously unsolved by ES. 109 of the index cases also received srGS not to miss SNVs and for determining the added value of lrGS, and 97 of the index cases received additional RNAseq of blood samples (n=80) or fibroblasts (n=17). In total, we solved 21 cases, of which 7 harbored complex SVs (n=5) or deep intronic variants (n=2) not detectable by ES in retrospect, while 14 were SNVs which were confirmed in ES re-analysis. All variants affected known epilepsy genes, and were identified in the initial lrGS analysis. Independent review of available srGS and/or RNA data did not reveal additional diagnoses. Identified (likely) pathogenic variants and their ACMG classification are listed in Table 1. We will first describe the 5 SVs including clinical features and a detailed comparison of the different sequencing methods, then the intronic variants, and last summarize the SNVs.

**Table 1:** Summary of (likely) pathogenic genetic variants identified with lrGS.

| ID | Gene | Variant type | Variant | Status | Inheritance | ACMG criteria* | Reason variant was missed in prior ES | Detection in ES/ srGS/lrGS |
|---|---|---|---|---|---|---|---|---|
| 1 | *CDKL5* | Translocation | t(X;17)(p22.13;p11.2), chrX:g.pter_18571502delins[chr17:g.pter_20929358], chr17:g.pter_20929358delins[chrX:g.pter_18571502] | heterozygous | *de novo* | 2E, 4C (P) | complex SV | srGS not available, lrGS |
| 2 | *SCN1A* | Inversion | chr2:164929688_166014987delins(AACT;164929688_166014984inv) | heterozygous | *de novo* | 2E, 4C (P) | complex SV | srGS, lrGS |
| 3 | *MEF2C* | Deletion | chr5:g.88872465_160652998delins[88883496_88885137;88916302_160616291inv] | heterozygous | *de novo* | 2E, 4C (P) | complex SV | srGS, lrGS (better characterization) |
| 4 | *STXBP1* | Deletion | chr9:127656273_127659122del | heterozygous | *de novo* | 2E, 4B (P) | complex SV mediated by transposons | srGS, lrGS (better characterization) |
| 5 | *CDKL5* | Deletion | c.745-400_825+1338delins[N[269]] | heterozygous | *de novo* | 2E, 4C (P) | complex SV mediated by transposons | Difficult to detect in srGS, lrGS (better characterization) |
| 6 | *SCN1A* | Intronic | c.265-2028C>G, p.?3 | heterozygous | *de novo* | PVS1, PM2. PS3 (P) | deep intronic variant | srGS, lrGS |
| 7 | *RNU2-2* | non-coding | n.[104T>G];[171_179] | compound-het | paternal; maternal | PS4, PP1, PM3 (LP) / PM1, PM2_supp, PM3, PP1 (LP) | non-coding gene | srGS, lrGS |
| 8 | *DEPDC5* | Deletion | chr22:31842375_31844024del | heterozygous | paternal | 2E (LP) | No CNV analysis in previous ES | ES, srGS, lrGS |
| 9 | *NR4A2* | Deletion | chr2:152816233_162128101del | heterozygous | *de novo* | 2E, 4C (P) | No CNV analysis in previous ES | ES, srGS, lrGS |
| 10 | *CDKL5* | Deletion | chrX:18409063_19203398del | heterozygous | *de novo* | 2E, 4C (P) | No CNV analysis in previous ES | ES, srGS, lrGS |
| 11 | *CDKL5* | Deletion | chrX:18575786_18586319del | heterozygous | *de novo* | 2E, 4C (P) | No CNV analysis in previous ES | ES, srGS, lrGS |
| 12 | *SCN8A* | Missense | c.653C>A, p.Ala218Glu | heterozygous | *de novo* | PM2, PP2, PP3, PS2, PS4 (LP) | No trio analysis in previous ES/*de novo* information | ES, srGS, lrGS |
| 13 | *TUBA1A* | Missense | c.518C>T, p.Pro173Leu | heterozygous | *de novo* | PP5, PM1, PP3, PM2, PS2 (LP) | clinical re-phenotyping | ES, srGS, lrGS |
| 14 | *NSF* | Missense | c.1688C>T, p.Pro563Leu | heterozygous | mosaic 23% in blood | PM5, PP3, PP5, PM2, PP2, PS2 (LP) | mosaicism calling | ES, srGS, lrGS |
| 15 | *FBXO11* | Missense | c.1781A>G, p.His594Arg | heterozygous | *de novo* | PP2, PP3, PS4, PS2 (LP) | updated gene panel / HPO match filter | ES, srGS, lrGS |
| 16 | *NR2F1* | Inframe deletion | c.298_300del, p.Gln100del | heterozygous | *de novo* | PM2, PM4, PS2, PP4 (LP) | No trio analysis in previous ES/*de novo* information, clinical re-phenotyping | ES, srGS, lrGS |
| 17 | *FZR1* | Missense | c.1333C>G, p.Arg445Gly | heterozygous | *de novo* | PP2, PP3, PM2, PS2 (LP) | No trio analysis in previous ES/*de novo* information | ES, srGS, lrGS |
| 18 | *CAMTA1* | Splice variant | c.4183-1G>A, p.? | heterozygous | *de novo* | PVS1, PS2, PM2 (P) | updated gene panel / HPO match filter | ES, srGS, lrGS |
| 19 | *RHOBTB2* | Missense | c.656C>T, p.Ser219Phe | mosaic 30% in blood | *de novo* | PP2, PP3, PM2, PS2 (LP) | mosaicism calling | ES, srGS, lrGS |
| 20 | *CHD2* | Frameshift | c.1652_1653del, p.Glu551ValfsTer2 | heterozygous | *de novo* | PVS1, PS2, PM2 (P) | updated gene panel / HPO match filter | ES, srGS, lrGS |
| 21 | *TMEM63B* | Missense | c.130G>A, p.Val44Met | heterozygous | *de novo* | PM2, PP2, PP3, PS2, PS4, PS3 (P) | updated gene panel / HPO match filter | ES, srGS, lrGS |

\*P = pathogenic, LP = likely pathogenic, ES = exome sequencing, srGS = short-read genome sequencing, lrGS = long-read genome sequencing

### Complex structural variants detected by lrGS and associated case reports

#### Balanced translocation leading to fusion transcript of *CDKL5*

A preschool-aged female (family 1) presented with focal tonic seizures that started in the first six months of life, which soon evolved to epileptic spasms. At the time of the last examination, she suffered from severe generalized muscular hypotonia, was non-verbal and non-ambulatory, had profound intellectual disability, and feeding difficulties. EEG showed diffuse slowing and multifocal epileptiform discharges. lrGS revealed a balanced *de novo* translocation t(X;17)(p22.13;p11.2). The patient had not previously undergone chromosomal analysis. Breakpoint evaluation in IGV revealed disruption of the *CDKL5* gene, with the breakpoint located within intron 4 (chrX:18571502) and a second breakpoint at chr17:20929358 (Fig. 1A). Fibroblast extracted RNAseq revealed the formation of a fusion transcript combining the first three exons of *CDKL5* fused with part of a lncRNA *CCDC144NL-AS1* downstream of the breakpoint on chr17, supporting loss of function of CDKL5. Additionally, RNAseq demonstrated extreme skewing of the X-chromosome in favor of expression of the fusion transcript, which was in line with the severe phenotype observed in the patient. Trio-based phasing showed that the disrupted allele originated from the paternal X chromosome. SrGS was not available for this patient. The translocation was later confirmed in a diagnostic setting using optical genome mapping.

**Figure 1.**
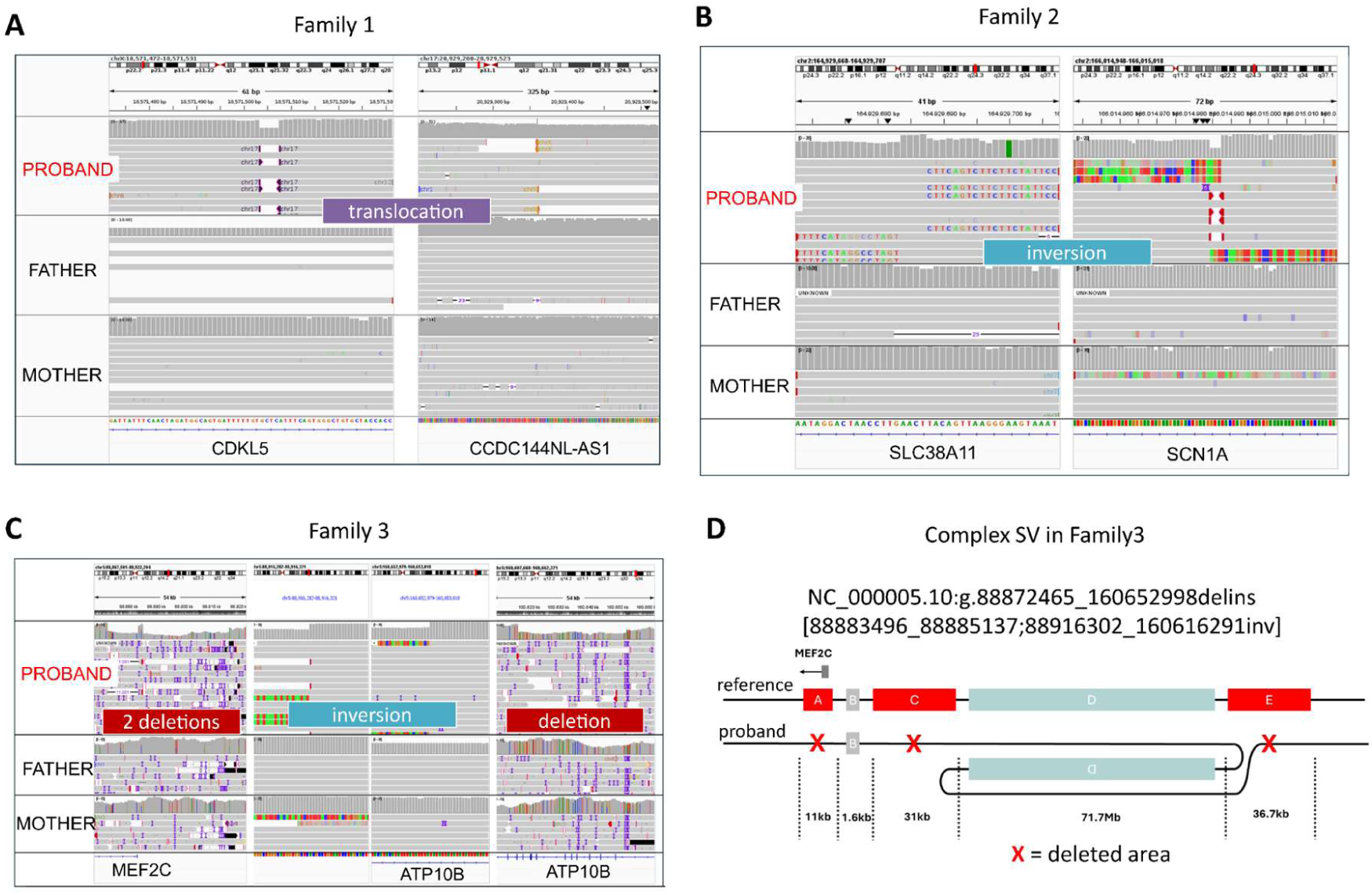
Complex structural variants. **A)** *De novo* balanced translocation disrupting transcript of *CDKL5* within intron 4 (chrX: 18571502) and chr17:20929358 leading to a fusion transcript with lncRNA *CCDC144NL-AS1* in Patient 1. **B)** Copy-neutral *de novo* inversion between intron 20 of *SCN1A* (chr2:166014984) and intron 7 of *SLC38A11* (chr2:164929688) accompanied by a 3 bp deletion in *SCN1A* intron 20 and a 4 bp duplication in *SLC38A11* in Patient 2. **C-D)** *De novo* 11,031 bp deletion of *MEF2C* first exon within a complex rearrangement comprising a large inversion (∼71Mb, chr5:88916302_160616291inv) and two deletions (chr5:88883496_88885137del, chr5:160616291_160652998del) in Patient 3.

#### Copy-neutral inversion between *SCN1A* and *SLC38A11*

An adolescent female (family 2) diagnosed with Dravet syndrome presented with fever-associated seizures in the first six months of life. Subsequently, she developed both fever-associated and afebrile seizures of multiple types (focal aware, generalized tonic-clonic, focal myoclonic to tonic and bilateral myoclonic seizures). She showed mild developmental delay with autistic features, crouched gait and mild scoliosis. EEG performed betweeen five and ten years revealed multifocal epileptiform discharges. lrGS revealed a *de novo* copy-neutral inversion with breakpoints in intron 20 of *SCN1A* (chr2:166014984) and intron 7 of *SLC38A11* (chr2:164929688). The rearrangement was accompanied by a 3 bp deletion at the *SCN1A* breakpoint and a 4 bp duplication at the *SLC38A11* breakpoint (Fig. 1B). RNAseq was inconclusive, since *SCN1A* is not expressed in blood nor fibroblasts. So far, *SLC38A11* has no established disease association, shows no intolerance to loss-of-function variation (pLI = 0, LOEUF = 1.17), indicating that loss of function is an unlikely disease-causing mechanism for this gene. The eight additional genes located within the inverted segment were not considered disrupted by the rearrangement. Upon subsequent srGS evaluation, the inversion and the breakpoints were well detectable and characterizable in singleton srGS as well, indicating that the diagnostic contribution in this case derived from genome rather than specifically long-read sequencing. *De novo* calling enabled rapid prioritization of this inversion and supported its interpretation.

#### Large chromosomal inversion with single exon deletion of *MEF2C*

A middle-aged woman (family 3) presented with severe global developmental delay starting in the first six months of life. She never acquired speech, and exhibited severe intellectual disability and autistic features. Motor development was mildly delayed. Epilepsy onset occurred between five and ten years of age with generalized tonic–clonic seizures, later evolving into pharmacoresistant epilepsy. EEG showed generalized epileptiform discharges and slow background activity. MRI showed mild cerebellar and parietal atrophy. Trio-lrGS identified a *de novo* 11,031 bp deletion affecting the first exon of *MEF2C* (chr5:88872465_88883496del) which was embedded within a complex rearrangement comprising two additional downstream deletions (chr5:88883496_88885137del and chr5:160616291_160652998del) and an approximately 71 Mb inverted insertion (chr5:88916302_160616291inv) between them (Fig. 1C, D). The single-exon *MEF2C* deletion was confirmed by qPCR, while the chromosomal analysis also confirmed the presence of a paracentric inversion of the long arm of chromosome 5 with breakpoints and rejoining points in bands q14 and q34, respectively. Expression analysis in blood-derived RNAseq showed reduced MEF2C expression (30%, p=0.001), consistent with partial loss of gene dosage. Subsequent srGS also identified the single-exon *MEF2C* deletion and the large inversion as separate events. Of note, srGS is associated with high false-positive rates for inversion calls and has particular difficulty resolving large inversions.^30^ Consequently, genuine large inversions may be dismissed as artefacts and overlooked.

#### Small Deletion of *STXBP1* with breakpoints in Alu-regions

An adolescent female (family 4) with severe global developmental delay, markedly limited speech (approximately 15 words), delayed independent walking with an ataxic gait, microcephaly, and a history of epileptic spasms with onset in the first six months of life, was found to carry a *de novo* heterozygous deletion at 9q33.3 (chr9:127656273_127659122del, GRCh38) encompassing exon 4 of *STXBP1*. This deletion was identified by trio-lrGS and was absent from gnomAD. This deletion was also called in srGS, however, due to the presence of flanking Alu elements, precise breakpoint resolution was not achievable using srGS, but with long-reads (Fig. 2A). Alignment analysis (Waterman-Eggert) of the flanking sequences ±100 bp of the reported breakpoints demonstrated >80% similarity of the immediate upstream reference sequence with the furthest downstream sequence of the deletion, and >88% similarity of the immediate downstream reference sequence with the furthest upstream sequence of the deletion. This is consistent with an established mechanism of deletion by recombination between homologous *Alu* elements.^31,32^ *STXBP1* is not expressed in blood-derived, precluding transcript-level evaluation.

**Figure 2.**
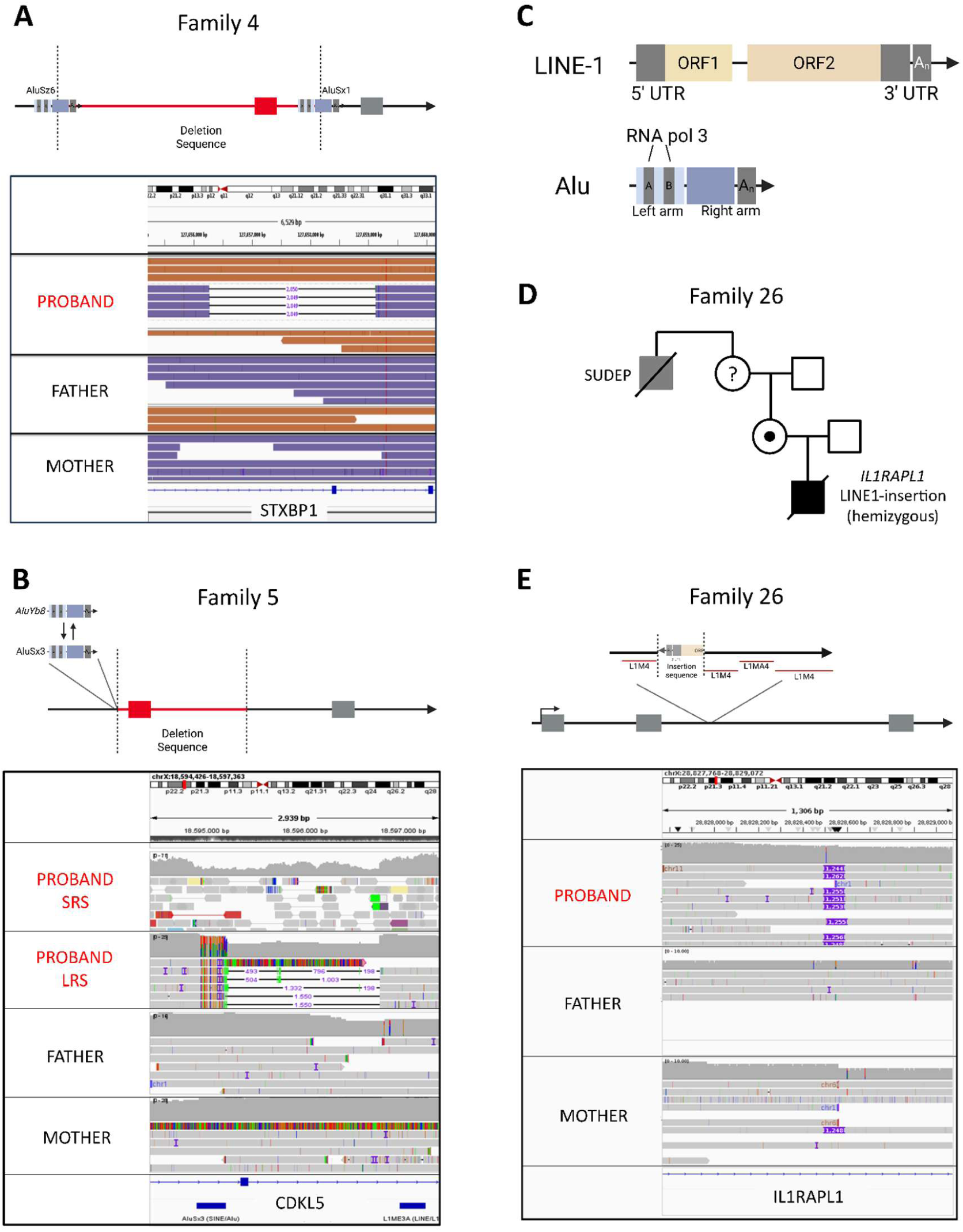
Complex structural variants mediated by transposons. **A)** *De novo STXBP1* deletion possibly mediated by non-allelic homologous recombination due to 98.1% overlap between *AluSz6* and *AluSx1* identified in Patient 4. **B)** *De novo Alu* insertion of *AluYb8* replacing *AluSx3* sequences followed by a 1.5kb deletion comprising exon 10 of *CDKL5* identified in Patient 5. **C)** Graphical Illustration of genomic arrangement of LINE-1 and Alu-elements. **D)** Pedigree of Family 26 (closed symbols indicate affected individuals, open symbols indicate healthy family members, carriers are marked with a dot within the symbol and possibly affected are colored grey). **E)** Hemizygous rare *LINE*-1 insertion within *IL1RAPL1* identified in Family 26, inherited from the mother and classified as VUS.

#### *Alu* retrotransposon-mediated exon deletion of *CDKL5*

Another female patient in her early teens (family 5) presented with tonic seizures starting in the first six months of life, which rapidly evolved into hypermotor-tonic-spasms sequence seizures, accompanied with hypsarrhythmia on EEG. Subsequently, she developed additional seizure types, including atypical absences and tonic seizures. At the time of the last follow-up, she had severe global developmental delay, lack of head control and profound intellectual disability, with hand stereotypies. Comorbidities included cortical visual impairment, sleeping problems and feeding difficulties. She was found to harbor a heterozygous deletion of exon 10 in *CDKL5* (Fig. 2B-C). Inspection in IGV revealed soft-clipped bases upstream of this deletion mapping to an *AluSx3* reference sequence. Using the Dfam database for transposable element sequence alignment, we traced the origin of these soft-clipped bases to a novel insertion of *AluYb8*, sharing approximately 87.6% sequence similarity with the original *AluSx3* element.^33^ The new *Alu* retrotransposon insertion resulted in exon-level deletion of *CDKL5*. Due to the mapping limitations of *Alu* elements in srGS, accurate breakpoint determination was only possible with lrGS. As *CDKL5* is not expressed in blood, and only blood-derived RNAseq data was available for this individual, transcript-level consequences could not be assessed.

### Causal variants missed in prior ES analyses or only detectable by GS

In addition to these complex SVs, causal CNVs were identified in four cases and causal SNVs in 12 cases; all likely pathogenic or pathogenic variants in epilepsy-related genes are listed in Table 1. In two cases, the identified SNVs were not detectable by ES, and required either sr- or lrGS: a deep-intronic *SCN1A* variant (c.265-2028C>G) identified in a patient (family 6) with Dravet syndrome (Fig. 3A), and bi-allelic non-coding variants in *RNU2-2* (family 7, n.[104T>G];[171_179]). The predicted inclusion of a 151-bp poison exon (c.265-2029_265-2179) caused by the *de novo* deep-intronic *SCN1A* variant was confirmed to be deleterious using a minigene assay (Fig. 3B-D).

**Figure 3.**
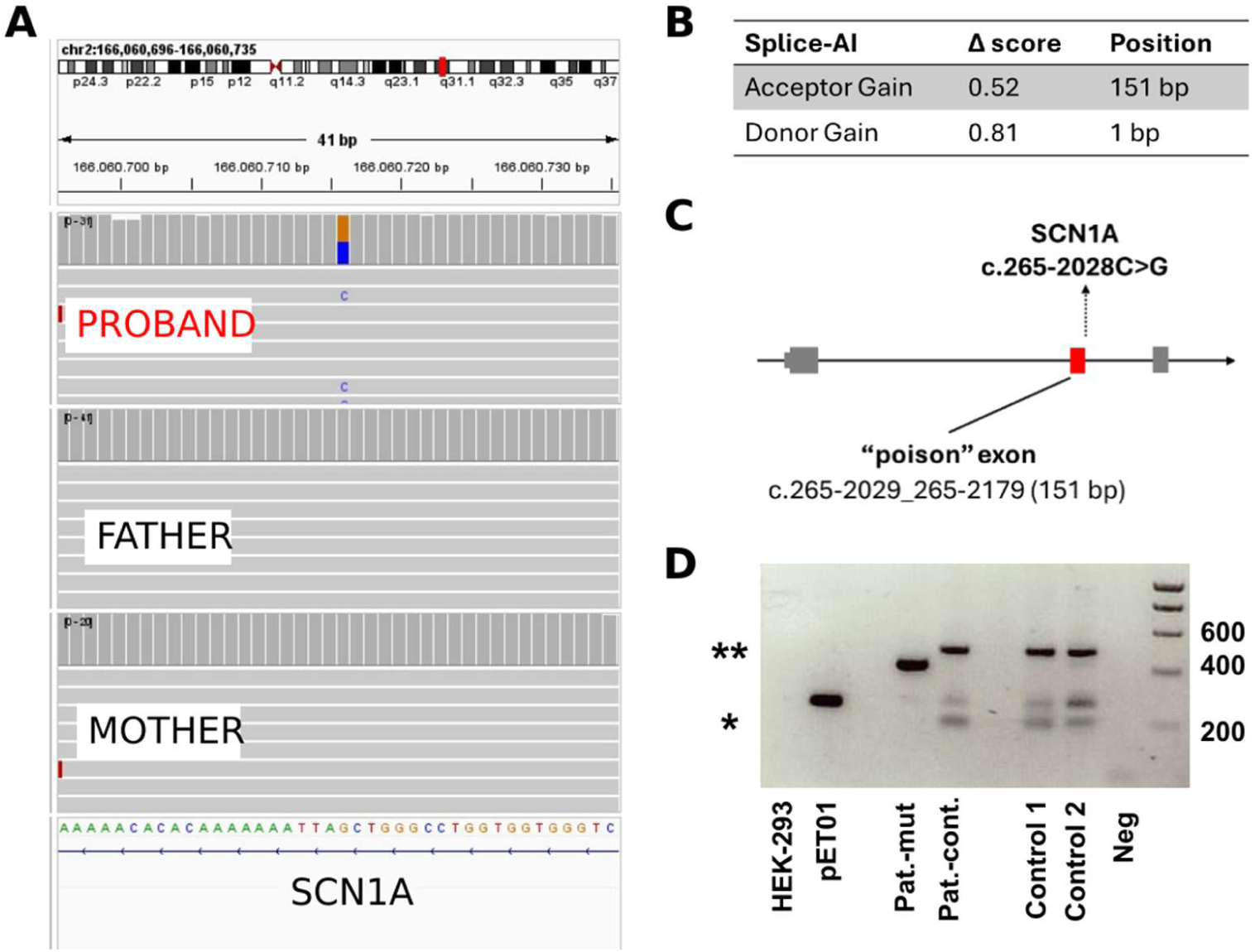
Cryptic exon inclusion mediated by *de novo* intronic variant. **A)** *De novo* intronic variant (c.265-2028C>G) detected in lrGS (Family 6). **B)** Splice-AI prediction of novel acceptor and donor site. **C)** Graphical illustration of the cryptic “poison” exon inclusion which possibly leads to a frameshift and nonsense-mediated decay. **D)** Mini-gene assay showed a dominant presence of the toxic exon (151 bp, sequence verified) in mRNA containing the intronic variant (Pat.-mut) compared to controls (Pat.-control, Control 1, Control 2). Interestingly, control samples also showed additional fragments besides the expected pET01 splice product. These fragments were subcloned and sequenced; one small fragment (*) was directly purified and sequenced. Its sequence indicated artificial splicing involving only rat insulin sequences from the pET01 vector. A second major fragment (**) contained a 228 bp alternative splice product corresponding to that described by Tang et al. (2025). Additionally, a 685 bp fragment was cloned and sequenced, representing another alternative splice product described by Tang et al. (2025), though it was only faintly visible on the gel.^61^

All CNVs and the other 10 SNVs were retrospectively detectable in prior ES but missed in the initial analysis. The CNV diagnoses (n=4) were previously missed due to absent CNV calling in local ES pipelines. Several pathogenic variants (n=4) were only detected after application of updated gene panels and HPO-based filtering, while some variants (n=3) were prioritized in this study by *de novo* calling and reverse phenotyping, since only singleton ES had been performed prior to the study for these families. These variants had previously been classified as variants of uncertain significance (VUS) and were not further investigated. Additional patients were diagnosed following improved mosaicism detection (n=2), or clinical re-phenotyping (n=1, Table 1). These findings demonstrate that a substantial proportion of diagnoses in apparently exome-negative DEE can be recovered through systematic reanalysis using updated analytical and interpretative approaches.

### Variants of Uncertain Significance in known epilepsy genes

Seven individuals harbored rare compound-heterozygous (n=5), hemizygous (n=1) and *de novo* heterozygous (n=1) variants in established epilepsy genes (*RNU2-2*, *IL1RAPL1, EPG5, ZNF335, TUBB3, LRP2*, and *SZT2*). Due to limited functional evidence and/or lack of phenotypic concordance, these variants were considered of uncertain, but possibly disease-relevant findings.

In the recently established disease gene *RNU2-2,* two previously unreported variants (n.73C>T; n.119G>A) were identified in a compound-heterozygous state and were classified as VUS.^34^ In a male patient with infantile epileptic spasm syndrome that later evolved to Lennox–Gastaut syndrome, a hemizygous intronic 1.2 kb insertion was identified in the X-Chromosome gene *IL1RAPL1*. The proband additionally showed severe intellectual disability with no speech development, independent walking for short distances and marked spasticity in the lower limbs. The inserted segment partially aligned with an *L1M4* subfamily element on the negative DNA strand. The patient’s mother was a heterozygous carrier, and the family history was suggestive of X-linked inheritance, including a maternal uncle who suffered from early childhood seizures and died in the first six months of life, possibly due to sudden unexpected death in epilepsy (SUDEP) (Fig. 2D-E). IL1RAPL1 is not expressed in blood and fibroblasts. As the patient had passed away between 35 and 40 years of age, no additional functional studies could be performed. Despite this suggestive family history, insufficient genetic evidence and the non-specific phenotype precluded definitive pathogenic classification. Compound-heterozygous variants in *SZT2* (c.5626T>C, p.Ser1876Pro; c.*1365C>T, p.?) were considered less likely to be causative, as blood-derived RNAseq revealed no significant changes in gene expression or splicing. In patient 23 harboring compound-heterozygous variants in *EPG5* (c.6724del, p.Met2242CysfsTer5; c.2718+945C>T, p.?) RNAseq showed only mildly reduced expression (17%, p=0.006). Similarly, a patient carrying compound-heterozygous variants in *LRP2* (c.13273G>A, p.Val4425Ile; c.2626G>T, p.Ala876Ser) displayed no features consistent with Donnai–Barrow syndrome, such as characteristic craniofacial, ocular, renal, or auditory findings. Following reevaluation, these variants were deemed unlikely to explain the phenotype. A rare *de novo* variant in *TUBB3* (c.643C>T, p.Leu215Phe) was identified in a patient with global developmental delay, complex brain malformation (partial corpus callosum dysgenesis, septum pellucidum agenesis, pontine atrophy, left hippocampus inversion and cerebellar malformation with partial vermis and hemisphere fusion) and pharmacoresistant structural epilepsy. Additional findings included bilateral optic nerve atrophy due to congenital hydrocephalus. Pathogenic *TUBB3* variants (OMIM #614039) are associated with midline brain malformations including vermis atrophy, consistent with this patient’s phenotype.^35^ Abnormal gyration has also been reported as a characteristic *TUBB3-*phenotype feature but this could not be assessed due to a lack of updated neuroimaging, as the family did not wish further investigations. Therefore, the variant remained classified as a VUS. Furthermore, a pathogenic *ZNF335* variant (c.3486_3487+24del) and a *de novo* missense variant (c.1519T>C, p.Trp507Arg) was detected in compound-heterozygous state in another individual, but insufficient clinical data - particularly regarding head circumference and neuroimaging - prevented definitive genotype-phenotype correlation.

### *ZNF569* and *ZNF570* identified as new candidate genes

In a young boy with Lennox-Gastaut syndrome (family 22), we identified a homozygous deletion affecting the first two exons of both *ZNF569* and *ZNF570*, two zinc-finger genes. This variant was not observed in the in-house databases, and observed only once in gnomAD v4 in a heterozygous state. GnomAD also contains no homozygous loss-of-function variants for either gene. Both parents were heterozygous carriers, and segregation analysis by PCR showed that the unaffected brother and sister were not homozygous for the deletion.

Both *ZNF569* and *ZNF570* are brain-expressed genes belonging to the large family of KRAB-containing C2H2 zinc-finger transcription factors, which play essential roles in neurodevelopment.^36^ Although population data suggests that both genes tolerate heterozygous loss, no homozygous loss-of-function variants were observed in gnomAD for either gene, suggesting that a complete loss may be deleterious. Notably, several related KRAB-ZNF genes are already known to be implicated in neurodevelopmental disorders and epilepsy, including *ZNF142*, in which biallelic variants cause a syndromic neurodevelopmental disorder with intellectual disability and early-onset seizures,^37^ and *ZNF711*, where pathogenic variants have been reported in individuals with intellectual disability and epileptic seizures.^38^ In the absence of additional affected individuals or functional evidence we considered this deletion a candidate locus requiring further validation.

### Epilepsy genes exhibit higher expression in fibroblasts than leukocytes

RNAseq contributed to variant interpretation in four families: In family 1, fibroblast-derived RNAseq confirmed a *CDKL5* fusion transcript, in family 3, blood-derived RNA showed mild reduced expression of *MEF2C* (30%, p=0.001), in family 8, blood-derived RNAseq demonstrated reduced *DEPDC5* expression and double exon skipping, consistent with a paternal deletion, and in family 9, blood-derived RNAseq showed 50% reduced expression of multiple genes consistent with the ∼9.3Mb *de novo* deletion encompassing *NR4A2*. The deletions in families 8 and 9 were simultaneously also prioritized via Solve-RD RNAseq project.^39^

Heterozygous deletions in *CDKL5* were identified in three patients (families 5, 10, and 11), including an *Alu*-mediated deletion. Their effects on expression could not be assessed by analysis of the blood-derived RNA that was available from these patients, whereas fibroblast RNAseq in the patient with a balanced translocation involving *CDKL5* (family 1) confirmed aberrant transcript expression. In family 4, fibroblast-derived RNAseq would also have provided transcript-level information on the identified *STXBP1* deletion, which was not captured in the blood-derived RNAseq. Our internal RNAseq database of PAX-blood (n=455) and fibroblast samples (n=129) revealed that 1,510 of 1,948 (77.5%) epilepsy genes are expressed in blood versus 1,669 of 1,948 in fibroblasts (85.7%; using a cutoff of average TPM>1).

## 4. Discussion

In this cohort of 124 individuals with DEE who remained undiagnosed after prior ES, a lrGS-based diagnostic workflow established a new molecular diagnosis in 16.9% (21/124). The majority of these diagnoses (14/21; 67%) were retrospectively detectable in the original ES data, whereas 7 (one third of the pathogenic variants (33%, 7/21), or 5,6% of the total cohort) were exclusively detectable in srGS and/or lrGS. In five cases the variants were complex SVs affecting established DEE genes. Although detectable with srGS, lrGS enabled complete reconstruction of the complex rearrangement, allowing accurate interpretation, particularly when repetitive regions such as mobile elements were involved. Our findings identify complex SVs as the variant class for which long-reads currently provided the most distinctive diagnostic information.

The high proportion of diagnoses retrospectively detectable in ES emphasizes the continuing value of systematic reanalysis in individuals with apparently negative genetic testing. Variants had been missed because of absent CNV calling, outdated virtual gene panels, lack of parental sequencing and de novo prioritization, limited sensitivity for mosaic variants, or conservative variant interpretation at the time of initial testing. Consequently, reanalysis of existing ES should remain an integral component of the diagnostic pathway before or alongside escalation to genome sequencing. The successful detection of these variants using lrGS nevertheless underscores its value as a robust orthogonal approach capable of detecting and validating a broad spectrum of variant types, extending beyond just complex SV detection.

lrGS performed particularly well in resolving SVs involving repetitive sequences. Two of the complex SVs identified in our study, the *CDKL5* exon 10 deletion (family 5) and the *STXBP1* deletion (family 4), were mediated by *Alu* elements (Fig. 2A, 2B). Furthermore, we detected a hemizygous LINE-1 insertion of uncertain significance in the X-linked *IL1RAPL1* gene in a male patient with DEE. Mobile elements retrotransposons constitute roughly 45% of the human genome and are well-recognized drivers of genomic instability through retrotransposition and non-allelic homologous recombination.^31,32,40^ Their repetitive nature complicates short-read alignment and breakpoint assignment. Long reads spanning the repetitive elements enabled precise breakpoint resolution and elucidation of the underlying rearrangement mechanism. Similarly, lrGS facilitated reconstruction of the *MEF2C* deletion and 71-Mb inversion as components of a single complex rearrangement. These cases illustrate that the value of lrGS extends beyond variant detection to establishing the architecture and mechanism of complex genomic events.

Our results also highlight one candidate locus warranting further investigation. The homozygous deletion affecting *ZNF569* and *ZNF570* (family 22) is of particular interest, as both genes encode zinc-finger transcription factors, a family increasingly implicated in neurodevelopmental regulation and transcriptional control.^36^ Complete loss of two adjacent regulatory brain-expressed genes provides a biologically plausible mechanism for neurodevelopmental phenotypes, including DEE. However, in the absence of functional data, this locus remains a preliminary candidate. Additional cases and experimental validation assessing their roles in neuronal development will be required to establish a definitive disease association.

Establishing a molecular diagnosis in DEE has increasing therapeutic relevance. We identified *SCN1A* disrupting variants in two individuals (family 2, family 6) with Dravet syndrome, one of the most extensively studied DEEs for which genotype-informed treatment strategies are already applied in the clinic.^41^ One of the *SCN1A* variants (index of family 6, deep-intronic *de novo SCN1A* variant c.265-2028C>G) results in a poison exon inclusion. Antisense oligonucleotides designed to suppress very similar poison exons are already being tested in phase 3 randomized controlled trials.^42^ Complex SVs were also identified in well-known genes such as *STXBP1* and *CDKL5*, for which targeted approaches are in development and supported by robust preclinical outcomes.^43–45^ Comprehensive identification and mechanistic characterisation of such SVs using lrGS may become increasingly relevant to identify patients who may benefit from emerging gene-targeted interventions.

Our analyses also highlight current technical challenges in clinical implementation of lrGS. Automated SV calling did not consistently capture the complete architecture of complex rearrangements, and manual review in integrative genomics viewer (IGV) was essential to identify additional breakpoints, correct orientation assignments, recognize mobile element involvement, or exclude alignment artifacts. For example, the *CDKL5* translocation (family 1) was initially reported as a head-to-head rearrangement; however, manual inspection demonstrated a head-to-tail orientation. Similarly, the *de novo MEF2C* deletion (family 3) was initially detected as a single-exon deletion. Careful visual inspection revealed it to be part of a complex rearrangement involving a large inversion and two additional deletions. Additionally, a substantial proportion of variants initially reported as *de novo* insertions in ONT data were, upon manual inspection, found to be alignment artifacts inherent to ONT technology rather than true insertions. To ensure consistency and reproducibility of variant detection across participating analysis centers, we performed cross-site validation using three positive controls drawn from the solved cohort described in this study (families 2, 3 and 5). This exercise revealed that even minor differences in bioinformatics pipelines, particularly software versions, could result in discrepant outcomes, either detecting or missing diagnostically relevant variants. Notably, one positive control (the *CDKL5 de novo* deletion) was missed when using Sniffles v2.6.1, but detected using an alternative workflow using Sniffles v2.4. These observations emphasize the need for standardized and validated analytical workflows, complementary SV callers, and careful visual review when lrGS is implemented diagnostically.

RNAseq provided functional evidence in four diagnosed individuals but did not independently identify additional diagnoses in this cohort. Its contribution was limited in part by tissue availability: most RNA samples were derived from blood, in which several relevant epilepsy genes are insufficiently expressed for meaningful transcript-level assessment. Our expression analysis indicates that fibroblasts provide broader coverage of epilepsy-associated genes than blood, and our previous work indeed showed that fibroblast-derived RNAseq significantly increases diagnostic yield in exome negative epilepsy patients.^46^

Several limitations should be considered. First, this was a clinically selected cohort rather than a population-based diagnostic study, and the observed yields may therefore not generalize to all individuals with ES-negative DEE. Second, sequencing platforms, coverage, and bioinformatic pipelines differed across subsets of the cohort. Third, srGS and RNA sequencing were not available for all participants, precluding a uniform head-to-head comparison of lrGS with these approaches across the entire cohort. Fourth, methylation information inherent to the long-read sequencing data was not analyzed because sufficiently validated and standardized analytical workflows were not available at the time of the study. This represents a potential source of additional diagnostic information: rare de novo differentially methylated regions have been reported to be enriched in individuals with unexplained neurodevelopmental disorders and can either provide diagnostic clues themselves or help identify underlying disease-causing (non-coding) genomic variants.^47,48^ Moreover, both PacBio HiFi and ONT sequencing have been shown to enable detection of disease-associated methylation signatures.^49,50^ Consequently, our study did not assess the potential incremental diagnostic contribution of methylation analysis, and therefore does not capture the full range of information that may ultimately be obtainable from a comprehensive lrGS-based diagnostic workflow.

In conclusion, trio lrGS contributed to molecular diagnosis in individuals with previously unresolved DEE, with its most distinctive value lying in the detection and structural resolution of complex rearrangements, particularly those involving repetitive and mobile-element sequences. At the same time, the finding that two-thirds of diagnoses were retrospectively detectable in prior ES highlights the substantial diagnostic value of systematic reanalysis. While lrGS greatly expands the spectrum of genomic variation that can technically be detected, the incremental diagnostic yields reported to date remain more modest than initially anticipated. Further advances may therefore depend increasingly on our ability to interpret variation outside the protein-coding genome. Recent discoveries of non-coding disease genes, including spliceosomal small nuclear RNA genes that account for a substantial proportion of previously unexplained NDDs,^7,34^ illustrate the diagnostic potential of this largely unexplored genomic space. Integrating sequence variation with additional information inherent to long-read data such as haplotype-resolved methylation profiles, may help prioritize and interpret non-coding variation and represents an important next step towards realizing the full diagnostic potential of lrGS.

## Materials and data availability

Genetic and phenotypic data for the majority of included individuals is accessible to registered users of the RD-Connect (<u>platform.rd-connect.eu</u>). Raw and processed data files generated in the Solve-RD project can be accessed through the European Genome-Phenome Archive (https://ega-archive.org) under study EGAS00001003851, following approval from the Solve-RD Data Access Committee.

## Ethics approval and consent to participate

This study was conducted according to the declaration of Helsinki and was approved by the institutional review board of Antwerp University Hospital (ethical committee number: 18/38/415) and by the ethics committee of University Hospital of Tübingen (ClinicalTrials.gov ID: NCT03491280). Informed consent for data sharing and publication was obtained from all participating individuals or their guardians.

## Acknowledgements

The authors thank the families and patients for participating in the study. LrGS was funded by the Federal Ministry for Research, Technology and Space (BMFTR, Treat-ION, grant #01GM2210A to H.L.), the German Research Foundation (DFG, 418081722 to T.B.H.), and the Research Foundation – Flanders (FWO, grants 1861424N and G056122N) and the Queen Elisabeth Medical Foundation (to S.W.). J.Pa. was supported by the Clinician Scientist program “PRECISE.net” funded by the Else Kröner-Fresenius-Stiftung and by the intramural TÜFF program (3049-0-0). J.K. was supported by the University of Tübingen (Fortuene Nr. 301100) and by the German Epilepsy Society (Otfrid-Foerster Stipend 2025), and is an associated member of Precise.net. Solve-RD has received funding from the European Union’s Horizon 2020 research and innovation programme under grant agreement No. 779257. This study makes use of data and tools shared/provided through the RD-Connect GPAP, which received funding originally from the European Union Seventh Framework Programme (FP7/2007-2013) under grant agreement No. 305444. The authors thank the Radboud Genome Technology Center for technical support. Part of the recruitment in German speaking countries was done with the help of the network ESNEK (Erhebung Seltener Neurologischer Erkrankungen im Kindesalter). This study was supported, in part, by funds from the ‘Current Research Annual Funding’ of the Italian Ministry of Health. This work was also supported by the Italian Ministry of Health, Ricerca Finalizzata (RF-2019-12370059 to D.M.). N.S. received funding from UA-Bijzonder Onderzoeksfonds (BOF)-DOCPRO4 (FFB180186). C.M. received funding from University of Antwerp-BOF (FFB200262). Z.E. received funding from the Marguerite-Marie Delacroix Support Fund, Belgium. E. C. was a recipient of the Marguerite-Marie Delacroix Support Fund Fellowship, Ghent, Belgium. This publication has emanated from research supported in part the Rare Diseases Research and Innovation Catalyst Awards (RDCat) 2023 from the Health Research Board of Ireland and by a research grant from Research Ireland under Grant Number 21/RC/10294_P2 and co-funded under the European Regional Development Fund and by FutureNeuro industry partners and by the Health Research Board. M.R.C. received research funding from the Fonds de Recherche Clinique, Saint-Luc University Hospital, Brussels, Belgium. Saint-Luc University Hospital is part of the Brussels Rare and Complex Epilepsies Consortium, a full member of the European Reference Network EpiCARE. J.G. and V.Y. was supported by the Deutsche Forschungsgemeinschaft (DFG, German Research Foundation) via the project NFDI 1/1 “GHGA - German Human Genome-Phenome Archive” (#441914366). S.O. received funding from the European Union via the project European Rare Disease Research Alliance (ERDERA), GA n°101156595, funded under call HORIZON-HLTH-2023-DISEASE-07 and from the German Research Foundation (DFG; research grant numbers 514060894, 514208594, 514177729, and OS 647/1-1). This study was supported, in part, by funds from the ‘Current Research Annual Funding’ of the Italian Ministry of Health, and the Italian Ministry of University and Research (MUR): #NEXTGENERATIONEU (NGEU) National Recovery and Resilience Plan (NRRP) project ‘A Multiscale integrated approach to the study of the nervous system in health and disease’ (MNESYS, PE0000006), assigned to S.B. and R.G.. T.B.H. received funding from the European Commission (Recon4IMD - GAP-101080997), the German Research Foundation (DFG; research grant numbers 418081722, 433158657, EJP-RD Artemis: 542553983), and the Clinician Scientist program “PRECISE.net 2.0” funded by the Else Kröner-Fresenius-Stiftung. Further funding to H.Le. was received from BMFTR (Treat-ION, grant #01GM2210A), and the DFG (FOR-2715, grants Le1030/16-2 and Le1030/23-1) and the foundation ‘no epilep’.

## Authors contributions

J.Pa., Z.E., C.V.-W., K.E., T.N., S.W., T.B.H., and H.L. conceptualized and designed the study. J.Pa., Z.E., J.K., D.M., N.S., R.L.-B., W.C., G.D., L.S., M.St., and S.O. performed genomic data analyses (srGS/lrGS), variant interpretation, and pipeline development. T.D.P., P.D.R. and M.Str performed lrGS analyses. C.V.-W., K.E., C.M., M.Sw., H.S., A.J., M.D.W., A.-S.S., F.B., S.B., L.C., E.C., B.C., E.F., J.K., P.K., R.K., L.L., R.M., H.M., B.A.N., O.M., E.M., M.P., J.Po., A.R., I.S., I.S.-A., D.V., C.W., M.W., C.W., L.S., A.S., L.Stro., R.G., G.L., S.B. contributed to clinical data collection, patient recruitment, and phenotyping. H.Loe. and T.O. conducted functional analysis (minigene assay). A.E.-C., J.G., and V.A.Y. performed Solve-RD RNAseq analysis. C.D., P.M., provided intellectual content for the curation of gene list. J.Pa, Z.E., J.K., and D.M. wrote and edited the manuscript. T.B.H., S.W., H.Le. provided supervision and substantially revised the manuscript. All authors reviewed and approved the final manuscript.

